# Variation in National COVID-19 Mortality Rates Across Asian Subgroups in the United States, 2020

**DOI:** 10.1101/2022.04.02.22273341

**Authors:** Jay J. Xu

## Abstract

Provisional U.S. national COVID-19 mortality data for the year 2020 analyzed by the CDC in March 2021 indicated that non-Hispanic Asians fared markedly better overall than other racial/ethnic minority groups–and marginally better than non-Hispanic Whites–in terms of age-adjusted mortality rates. However, Asians in the United States are composed of diverse array of origin subgroups with highly varying social, economic, and environmental experiences, which influence health outcomes. As such, lumping all Asians together into a single category can mask meaningful health disparities among more vulnerable Asian subgroups. To date, there has not been a national-level analysis of COVID-19 mortality outcomes between Asian subgroups. Utilizing final multiple cause of death data for 2020 and population projections from the U.S. Census Bureau’s Current Population Survey Annual Social and Economic Supplement for 2020, crude and age-adjusted national COVID-19 mortality rates, both overall and stratified by sex, were calculated for the six major single-race Asian origin subgroups (Asian Indian, Chinese, Filipino, Japanese, Korean, and Vietnamese) and a catch-all seventh category that comprises the remaining Asian subgroups (Other Asians), contrasting them to the corresponding mortality rates of other racial/ethnic groups. A substantially more nuanced picture emerges when disaggregating Asians into its diverse origin subgroups and stratifying by sex, with Filipino males and Asian males outside of the six major Asian subgroups in particular experiencing markedly higher age-adjusted mortality rates than their White male counterparts, whether comparisons were restricted to their non-Hispanic subsets or not. During the COVID-19 pandemic and in the post-pandemic recovery, it is imperative not to overlook the health needs of vulnerable Asian populations. Public health strategies to mitigate the effects of COVID-19 must avoid viewing Asians as a monolithic entity and recognize the heterogeneous risk profiles within the U.S. Asian population.

## Introduction

Provisional U.S. national COVID-19 mortality data for the year 2020 analyzed by the Centers for Disease Control and Prevention (CDC) in March 2021 indicated that non-Hispanic (NH) Asians as a whole fared markedly better than other racial/ethnic minority groups–and marginally better than NH Whites–in terms of age-adjusted mortality rates during the first calendar year of the COVID-19 pandemic [1]. While the Asian population is relatively small, with the single-race segment of the Asian population comprising approximately 6% of the U.S. population, it is far from a monolith. Tracing ancestral lineages across the vast continent of Asia, the U.S. Asian population is highly diverse across multiple dimensions, varying considerably in spoken languages, cultural practices, historical pathways to America, geographical patterns of settlement, the degree of assimilation into mainstream American life and culture, and socioeconomic profiles [2]. For example, the substantial heterogeneity within the Asian population is reflected in part by its wide income inequality [3], highest among all racial groups in 2016 (as measured by the Gini coefficient) [4], which is robustly linked to health disparities [5, 6].

Although data aggregation is a common practice to increase sample size and statistical power to detect differences between population subgroups, lumping data together under the broader Asian umbrella term can mask meaningful health disparities among more vulnerable Asian subgroups, a phenomenon that is well-documented in the health sciences [7–27]. Disaggregated data on COVID-19 outcomes for different Asian subgroups have been largely absent throughout the pandemic [28, 29], and provisional datasets on population-level COVID-19 outcomes disaggregated by race that were publicly released by the Centers for Disease Control and Prevention (CDC) throughout 2020 and 2021 typically either included Asians as a single overall group or even combined them with Pacific Islanders.

While limited research has uncovered disparities in COVID-19 mortality outcomes among specific Asian subgroups in specific geographic regions and contexts, such as Hmongs in Minnesota [30], Chinese and South Asians in New York City [31], and Filipinos in California [32], little to no attention by national media outlets and public health officials has been given to vulnerable Asian subgroups [33]. To date, there has not been a national-level analysis of COVID-19 mortality outcomes between Asian subgroups in the U.S. To this end, I utilized final multiple cause of death data for 2020 that were released in December 2021 and population projections from the U.S. Census Bureau’s Current Population Survey Annual Social and Economic Supplement ASEC for 2020 to calculate crude, age-adjusted, and sex-stratified national COVID-19 mortality rates for the six major Asian origin subgroups (Asian Indians, Chinese, Filipino, Japanese, Korean, Vietnamese), as well as a catch-all seventh category that comprises the remaining less-populous Asian subgroups (Other Asians), contrasting them with the corresponding mortality rates of other racial/ethnic groups.

## Materials and methods

### Data sources

Final multiple cause of death data for 2020 were released in December 2021 [34], which are publicly available to download in machine readable format from the CDC WONDER online data portal [35]. At the time of writing, mortality data beginning in 2021 was considered provisional. From the CDC WONDER online data portal, national COVID-19 death counts (International Classification of Diseases, 10^th^ Revision code U07.1 as the underlying cause of death) jointly stratified by size-15 single-race category, Hispanic/Latino (Hispanic) ethnicity, sex, and single-year age group for the year 2020 were extracted. Separately extracted were COVID-19 death counts by size-15 single-race category for each state; death counts less than 10 for the state-level data were suppressed to protect confidentiality. In 2020, a total of 350,861 COVID-19 deaths occurred among U.S. residents, of which 1,427 (0.4%) deaths had unknown Hispanic ethnicity, and 4 deaths had unknown age. Among the 1,427 COVID-19 deaths in 2020 with unknown Hispanic ethnicity, 867 (60.8%) occurred among Whites, 460 (32.2%) occurred among Blacks, 82 (5.7%) occurred among Asians, 14 (1.0%) occurred among American Indian and Alaska Natives (AIAN), and 4 (0.3%) occurred among persons of two or more races.

Population projections from the U.S. Census Bureau’s Current Population Survey (CPS) Annual Social and Economic Supplement (ASEC) Public Use Microdata Sample data file for 2020, obtained from the IPUMS-CPS database [36], were used to obtain the denominators for all mortality rate calculations and to compute demographic summary statistics. Ages in the CPS ASEC data were binned into 5-year age intervals up until age 80 (i.e., ages 0–4, 5–9,…, 75–79, ≥ 80).

### Statistical analysis

The following single-race categories were considered: Whites, Blacks, Pacific Islanders, American Indian and Alaska Natives (AIAN), the six major single-race Asian origin subgroups (Asian Indian, Chinese, Filipino, Japanese, Korean, and Vietnamese), and a catch-all seventh Asian subgroup category that comprises the remaining less-populous Asian subgroups, termed Other Asians. Additionally, a single overall Asian racial group that combined all of the Asian subgroups was considered. Following the standard approach used by the CDC and in health disparities research more broadly, mortality rate comparisons were made between the NH subsets of the examined single-race categories as well as a racial/ethnic category comprising persons of Hispanic ethnicity, who can be of any race (i.e., Hispanics). Crude and age-adjusted mortality rates were calculated for each racial/ethnic group, both overall and stratified by sex, and age-adjusted mortality rates were calculated using direct age adjustment with respect to the 2000 U.S. Standard Population [37–39] after appropriately binning ages in the mortality data and standard population to align them with the age groups used for the CPS ASEC data. In addition, age-adjusted and sex-stratified mortality rates for NH Asians as a whole, each examined NH Asian subgroup, and NH Whites were calculated for each of the four following age categories: (i) ages < 45, (ii) ages 45–64, (iii) ages ≥ 65, and (iv) ages ≥ 75. The corresponding age-adjusted and sex-stratified mortality rate ratios and mortality rate differences for NH Asians as a whole and for each examined NH Asian subgroup were subsequently calculated, anchoring comparisons to NH Whites. To evaluate the robustness of comparisons between the examined racial groups without restricting comparisons to their NH subsets, a supplementary analysis was performed that first collapsed the mortality data over Hispanic ethnicity before calculating quantities analogous to those calculated the primary analysis.

The number of COVID-19 deaths with unknown Hispanic ethnicity (1,427) constitute less than half of a percent of total COVID-19 deaths and were excluded for the calculation of crude mortality rates in the primary analysis. Likewise, the 1,431 deaths with missing Hispanic ethnicity or age were excluded for the calculation of age-adjusted mortality rates in the primary analysis. In the supplementary analysis, no deaths were excluded for the calculation of crude mortality rates, and only the four deaths with missing ages were excluded for the calculation of age-adjusted mortality rates.

## Results

There were a total of 12,884 (single-race) Asian COVID-19 deaths in 2020, representing 3.7% of the total U.S. COVID-19 death toll for 2020. Table 1 displays the distribution of these deaths by Asian subgroup, contrasting their share of total Asian COVID-19 deaths with their share of the U.S. population. Nearly six in ten (59.2%) Asian COVID-19 deaths occurred among males, despite Asian males comprising less than half (47.8%) of the Asian population. The greatest discrepancies between their share of COVID-19 deaths and their share of the Asian population were observed for Filipinos and Asian Indians. Filipinos comprised the greatest share of Asian COVID-19 deaths among all the examined Asian subgroups (23.1%), over 80% higher than the share of Filipinos in the Asian population (12.7%). The disparity was greater among Filipino males, with their share of COVID-19 deaths nearly 2.5 times greater than their share of the Asian population (13.3% vs. 5.4%). In contrast, the Asian Indian share of Asian COVID-19 deaths was approximately half of the Asian Indian share of the Asian population (13.9% vs. 27.0%), and the disparity was greater among females, with their share of Asian COVID-19 deaths approximately equal to one-third of their share of the Asian population (4.3% vs. 12.9%). Over two-thirds (67.1%) of all Asian COVID-19 deaths occurred in just five states: California, Illinois, New Jersey, New York, and Texas.

**Table 1.** Number of COVID-19 Deaths, Share of Asian COVID-19 Deaths, and Share of the Asian Population by Asian Subgroup.

| Sex | Asian Subgroup | Deaths | Share of Asian Deaths | Share of Asian Population |
| --- | --- | --- | --- | --- |
| Overall | Asian Indian | 1,788 | 13.9% | 27.0% |
|  | Chinese | 2,613 | 20.3% | 20.0% |
|  | Filipino | 2,982 | 23.1% | 12.7% |
|  | Japanese | 611 | 4.7% | 4.1% |
|  | Korean | 1,159 | 9.0% | 7.9% |
|  | Vietnamese | 1,330 | 10.3% | 9.2% |
|  | Other Asian | 2,401 | 18.6% | 19.0% |
|  | **Total** | **12,884** | **100%** | **100%** |
| Male | Asian Indian | 1,229 | 9.5% | 14.1% |
|  | Chinese | 1,499 | 11.6% | 9.4% |
|  | Filipino | 1,718 | 13.3% | 5.4% |
|  | Japanese | 237 | 1.8% | 1.6% |
|  | Korean | 554 | 4.3% | 3.7% |
|  | Vietnamese | 833 | 6.5% | 4.1% |
|  | Other Asian | 1,553 | 12.1% | 9.5% |
|  | **Total** | **7,623** | **59.2%** | **47.8%** |
| Female | Asian Indian | 559 | 4.3% | 12.9% |
|  | Chinese | 1,114 | 8.7% | 10.7% |
|  | Filipino | 1,264 | 9.8% | 7.3% |
|  | Japanese | 374 | 2.9% | 2.5% |
|  | Korean | 605 | 4.7% | 4.2% |
|  | Vietnamese | 497 | 3.9% | 5.0% |
|  | Other Asian | 848 | 6.6% | 9.5% |
|  | **Total** | **5,261** | **40.8%** | **52.2%** |

Figure 1 displays the crude mortality rates for each racial/ethnic group in the primary analysis, both overall and stratified by sex. Crude mortality rates for NH Asians as a whole and for every NH Asian subgroup, with the exception of NH Filipinos, were lower than those of NH Whites, NH Blacks, Hispanics, and NH AIAN’s, both overall and stratified by sex. Overall, NH Filipinos had the third-highest crude mortality rate among all examined racial/ethnic groups behind NH Blacks and NH AIAN’s, and among males, NH Filipinos had the second-highest crude mortality rate behind only NH AIAN’s. The NH Filipino crude mortality rate was 39.4% higher than that of NH Whites among males, but it was 13.2% lower than NH Whites among females. The crude mortality rate for males was 58.0% higher than that of females among NH Asians as a whole, but the sex gap in crude mortality rates varied widely by NH Asian subgroup. While the male and female crude mortality rates for NH Japanese and NH Koreans were approximately equal, the crude mortality rates for males were over twice that of females among NH Asian Indians and NH Vietnamese.

**Figure 1.**
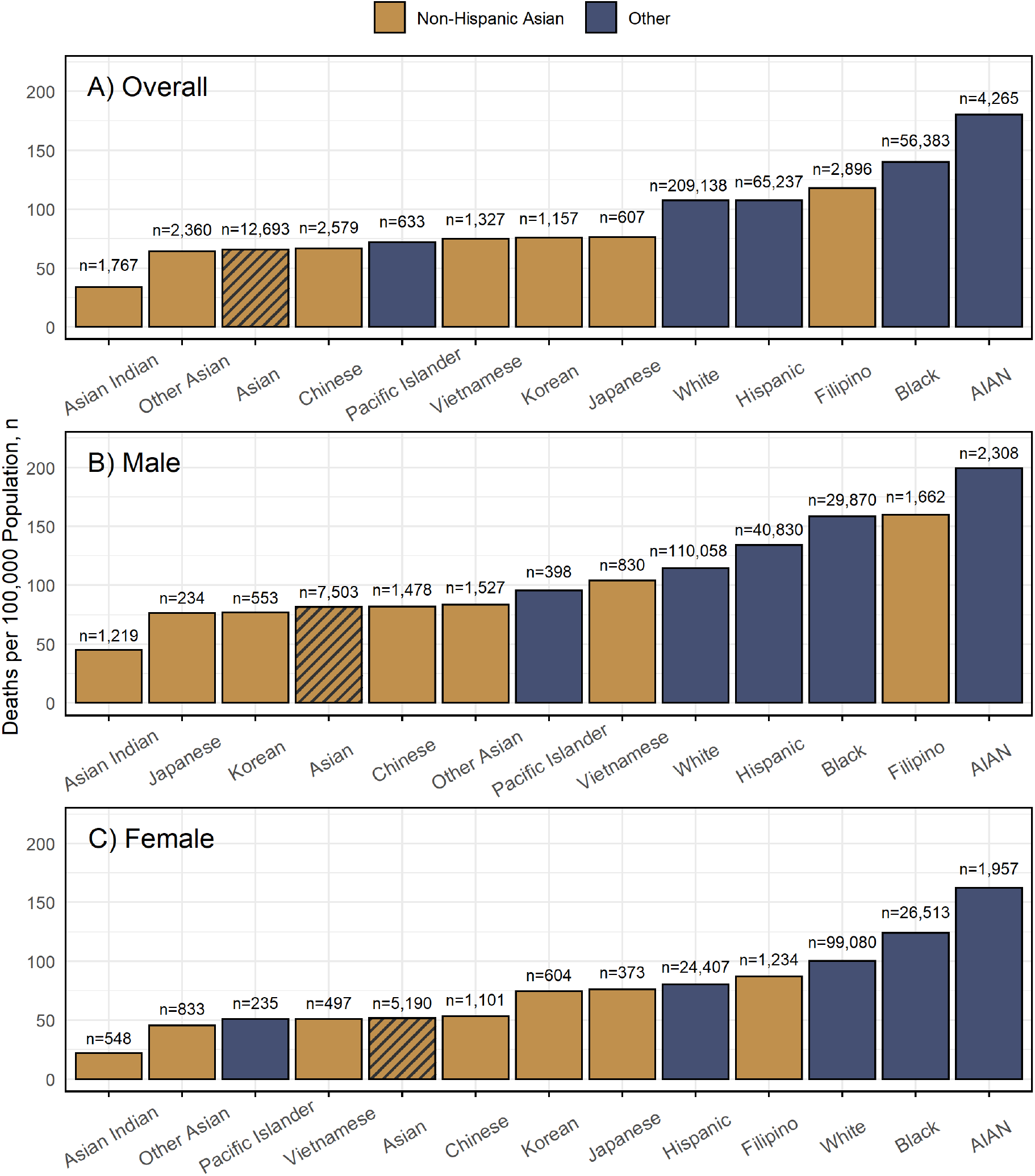
National Crude COVID-19 Mortality Rates by Race/Ethnicity in 2020.

Figure 2 displays the age-adjusted mortality rates for each racial/ethnic group in the primary analysis, both overall and stratified by sex. File S1 contains the age-adjusted and sex-stratified mortality rates for NH Asians as a whole, the seven NH Asian subgroups, and NH Whites for ages <45 years, ages 45–64 years, ages ≥ 65 years, ages ≥ 75 years, and all ages, and the corresponding mortality rate ratios and mortality rate differences relative to NH Whites. Age-adjusted mortality rates for NH Asians as a whole and for every NH Asian subgroup, with the exception of NH Other Asians, were smaller than those of Hispanics, NH Blacks, and NH AIAN’s, both overall and stratified by sex. Among males, the age-adjusted mortality rate for NH Asians as a whole was higher than that of NH Whites by 5.7%, a disparity that was driven by younger age groups; in fact, age-adjusted mortality rates for NH White men aged ≥ 65 and ≥ 75 exceeded that of NH Asian men (Table S1.1 in File S1). However, the age-adjusted mortality rate for NH Asians as a whole was lower among females by 16.1%, resulting in an age-adjusted mortality rate for NH Asians as a whole that was 5.3% lower than that of NH Whites overall.

**Figure 2.**
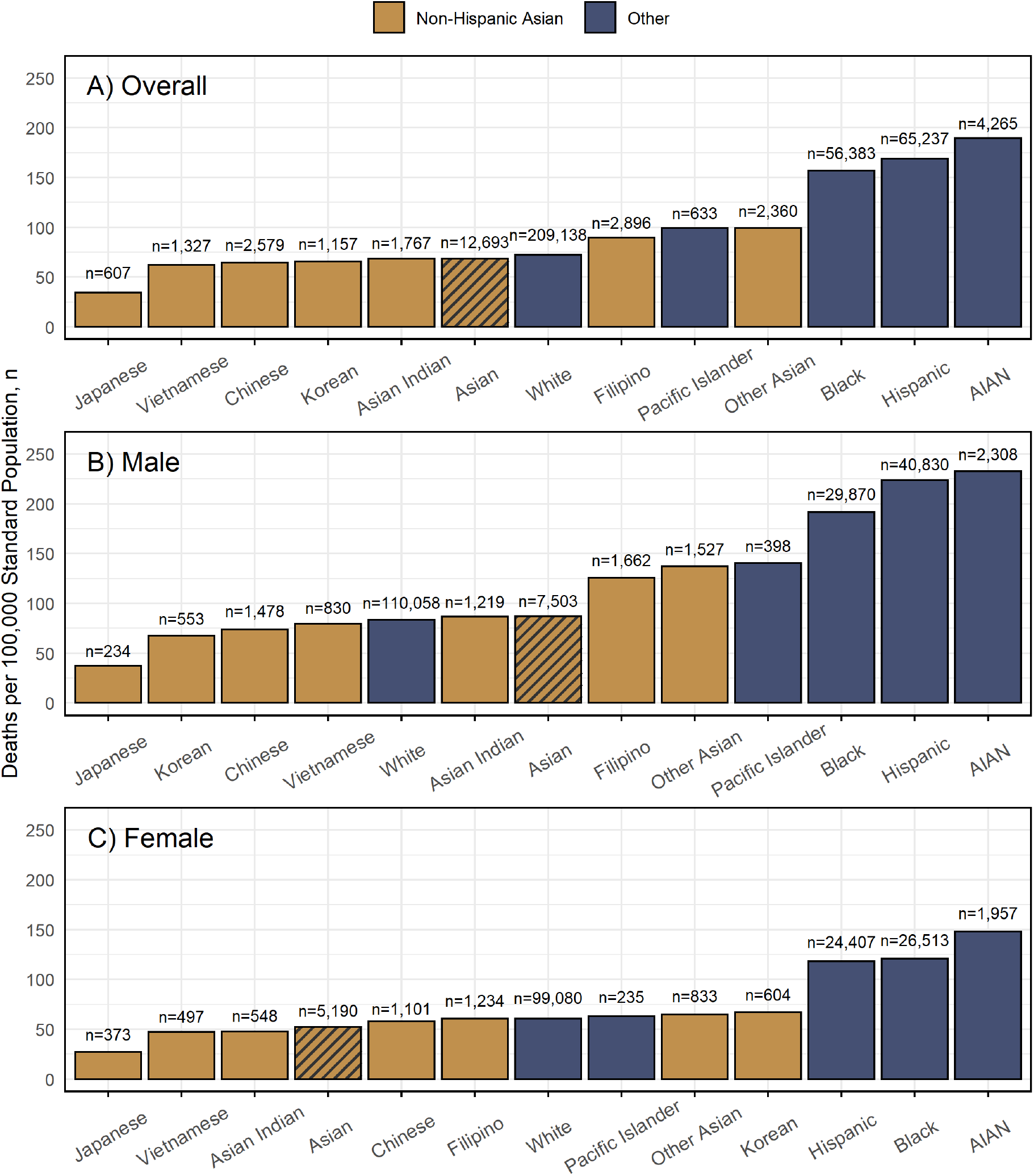
National Age-Adjusted COVID-19 Mortality Rates by Race/Ethnicity in 2020.

The age-adjusted mortality rate for NH Filipinos and NH Other Asians were higher than that of NH Whites overall, a disparity that was driven by males in younger age groups (Tables S1.4 and S1.8 in File S1). Among NH Filipinos, the age-adjusted mortality rate was approximately one and a half times higher among males but was approximately equal among females, and among NH Other Asians, the age-adjusted mortality rate was nearly two-thirds (64.1%) higher among males but was only 6.6% higher among females. While age-adjusted mortality rates for NH Koreans and NH Asian Indians were below that of NH Whites overall, directionally opposite trends were observed when stratified by sex. For NH Koreans, the age-adjusted mortality rate was 10.4% higher than NH Whites among females but was 19.4% lower among males. For NH Asian Indians, the age-adjusted mortality rate was 20.9% lower than NH Whites among females but was 3.4% higher among males. Age-adjusted mortality rates for NH Chinese, NH Japanese, and NH Vietnamese were below that of NH Whites, both overall and stratified by sex. The age-adjusted mortality rate for males was approximately two-thirds higher than that of females among NH Asians as a whole, but the sex gap in age-adjusted mortality rates varied widely by NH Asian subgroup. While the male and female age-adjusted mortality rates for NH Koreans were approximately equal, the age-adjusted mortality rates for males were over twice that of females among NH Filipinos and NH Other Asians.

Figure 3 displays the crude mortality rates for each examined racial group in the supplementary analysis, both overall and stratified by sex. Crude mortality rates for Asians as a whole and for every Asian subgroup, with the exception of Filipinos, were lower than those of Whites, Blacks, and AIAN’s, both overall and stratified by sex. Overall, Filipinos had the third-highest crude mortality rate among all examined racial groups behind Blacks and AIAN’s, and among males, Filipinos had the second-highest crude mortality rate behind only Blacks. While the Filipino crude mortality rate was 23.1% higher than that of Whites among males, it was 13.0% lower than Whites among females. The crude mortality rate for males was 58.0% higher than that of females among Asians as a whole, but the sex gap in crude mortality rates varied widely by Asian subgroup. Among Japanese, the crude mortality rate was in fact lower among males than females, and among Koreans, the crude mortality rate was only 5.4% higher among males. In contrast, the crude mortality rates for males were over twice that of females among Asian Indians and Vietnamese.

**Figure 3.**
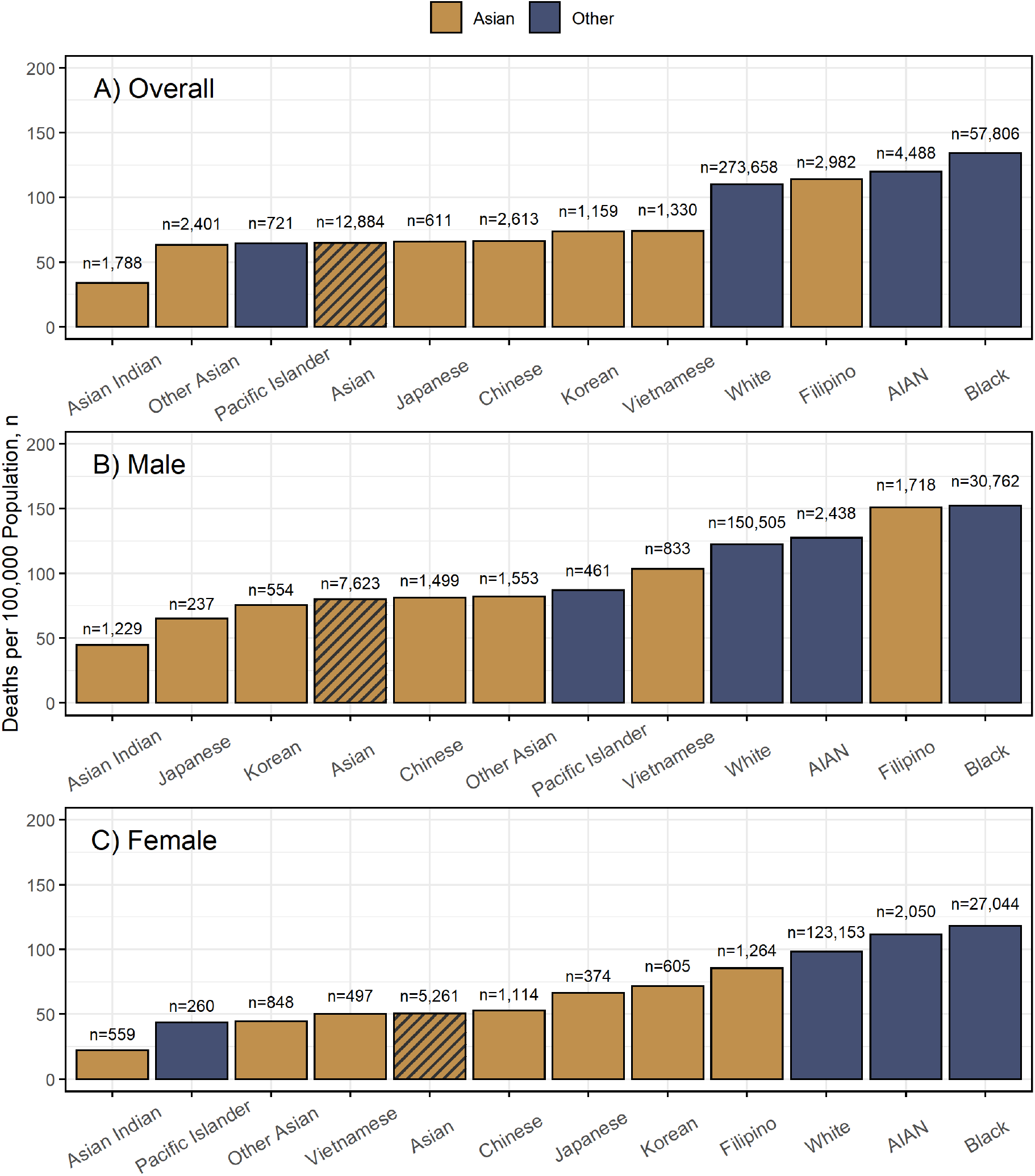
National Crude COVID-19 Mortality Rates by Race in 2020.

Figure 4 displays the age-adjusted mortality rates for each racial group in the supplementary analysis, both overall and stratified by sex. File S2 contains analogous quantities as File S1 after collapsing the mortality data over Hispanic ethnicity. Age-adjusted mortality rates for Asians as a whole were below those of all of the examined non-Asian racial groups, both overall and stratified by sex. Moreover, age-adjusted mortality rates for every Asian subgroup, with the exception of Filipinos and Other Asians, were below those of Whites, Blacks, and AIAN’s, both overall and stratified by sex. Age-adjusted mortality rates for Filipino and Other Asians exceeded that of Whites overall, a disparity that was driven by males below the age of 75 (Tables S2.4 and S2.8 in File S2). Among males, age-adjusted mortality rates for Filipinos and Other Asians were higher than that of Whites by 24.3% and 29.8%, respectively, but among females, age-adjusted mortality rates for Filipinos and Other Asians were below that of Whites by 12.9% and 9.3%, respectively. Among Asians as a whole, the age-adjusted mortality rate for males was approximately two-thirds higher than that of females, but the sex gap in age-adjusted mortality rates varied widely by Asian subgroup. While the male and female age-adjusted mortality rates for Koreans were approximately equal, the age-adjusted mortality rates for males were over twice that of females among Filipinos and Other Asians.

**Figure 4.**
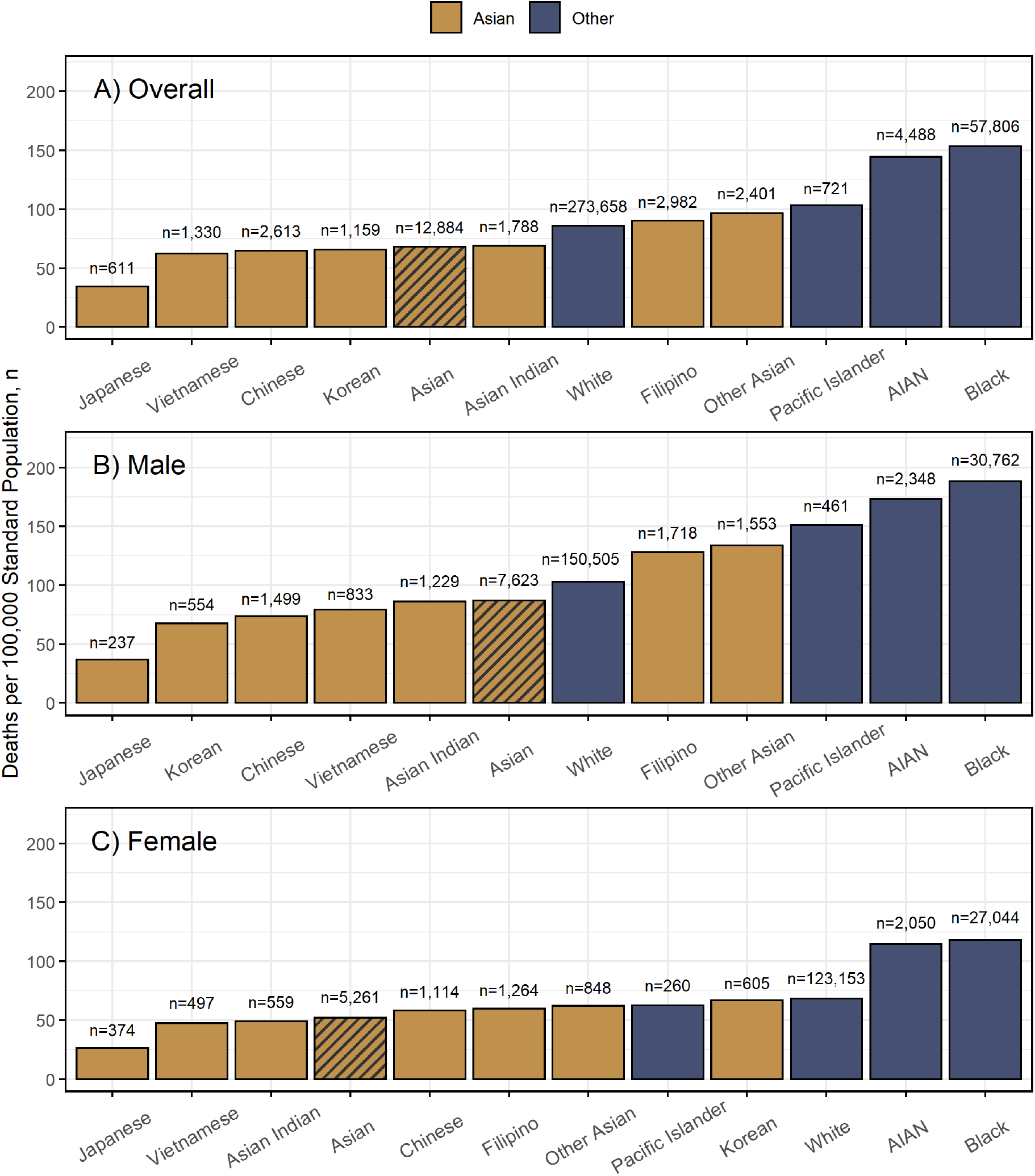
National Age-Adjusted COVID-19 Mortality Rates by Race in 2020.

## Discussion

This is the first national analysis of COVID-19 mortality rates for different Asian subgroups in the U.S. The results of the current analysis are in alignment with the aforementioned March 2021 CDC provisional analysis of COVID-19 deaths in 2020 which indicated that, as a single category, NH Asians fared markedly better than other racial/ethnic minority groups overall–and marginally better than NH Whites–in terms of age-adjusted mortality rates during the first calendar year of the COVID-19 pandemic. However, when Asians are disaggregated into subgroups and stratified by sex, a substantially more nuanced picture emerges. Chinese, Japanese, and Vietnamese Asian subgroups experienced lower COVID-19 mortality rates in 2020 than Whites, Blacks, Hispanics, and AIAN’s among both males and females, whether mortality rates were age-standardized or not and whether racial comparisons were restricted to their NH subsets or not. In contrast, mortality rates for Filipino males exceeded that of White males whether mortality rates were age-standardized or not and whether the Filipino-White comparison was restricted to their NH subsets or not. Moreover, the age-adjusted mortality rate for Other Asian males was higher than that of White males whether the comparison was restricted to their NH subsets or not. Age-adjusted mortality rates for NH Asian Indian males also exceeded that of NH White males, and age-adjusted mortality rates for NH Korean and NH Other Asian females were above that of NH White females.

While Asians as a whole had a lower age-adjusted mortality rate than Whites overall, whether the Asian-White comparison was restricted to their NH subsets or not, the gap was larger when the Asian-White comparison was not restricted to their NH subsets. This is presumably because single-race Whites comprise a substantially higher share of the Hispanic population than single-race Asians (88.2% vs. 1.0%), and Hispanic Whites are a higher-risk group than NH Whites, thereby inflating the White age-adjusted mortality rate while leaving the Asian age-adjusted mortality rate largely unchanged after including decedents of Hispanic ethnicity in the Asian-White comparison. A qualitatively similar pattern was also observed among Asian and White females. Among males, however, the NH Asian age-adjusted mortality rate in fact exceeded that of NH Whites, but the age-adjusted mortality rate for Asian males fell below that of White males after including decedents of Hispanic ethnicity in the Asian-White comparison. The Filipino-White and Other Asian-White age-adjusted mortality rate gaps among males attenuated after the inclusion of Hispanic decedents in their respective comparisons with Whites, but they did not close.

Limitations of the current analysis include the possibility of COVID-19 deaths that occurred in 2020 among whom COVID-19 went undiagnosed, as well as the possibility of racial/ethnic misclassification of COVID-19 deaths on official death certificates. A 2016 CDC study estimated a race misclassification rate of 3% on death certificates for Asians [40]. The same report, however, estimated race misclassification for AIAN’s to be 40%, which has been a long-standing and persistent problem [41–48]. Asian subgroup misclassification on death certificates among Asian decedents is also a potential limitation, but its current extent is unclear. Moreover, the current analysis is based on COVID-19 deaths that occurred in 2020, at the tail end of which the U.S. vaccination campaign began (December 14) [49]. As such, racial/ethnic patterns in mortality outcomes observed in the current analysis may not necessarily reflect patterns that unfolded during later stages of the COVID-19 pandemic in 2021 and 2022 when COVID-19 vaccines became widely available and when the more transmissible and virulent Alpha, Delta, and Omicron SARS-CoV-2 variants emerged and successively usurped previously dominant variants. All mortality rates presented in the current analysis also contain the caveat that the denominators used to calculate them are based on population projections derived from a national survey, namely the CPS ASEC, which in fact experienced significant operational challenges in 2020 as a result of the COVID-19 pandemic [50, 51]. The CPS ASEC was used as the data source for all mortality rate denominators so that mortality rates for all of the examined racial groups were calculated in a consistent manner that used a common data source, despite U.S. Census Bureau intercensal population estimates for 2020 being available for the non-Asian subgroup race categories examined [52], which are presumably more accurate. In short, mortality rates presented here should be interpreted with a moderate degree of caution.

Underlying reasons for the wide variation in COVID-19 mortality rates across Asian subgroups likely include differential rates of comorbid conditions in conjunction with a complex web of social, economic, and environmental factors. For example, across the six major Asian subgroups, Filipinos experienced the highest crude and age-adjusted mortality rates overall in 2020, whether comparisons were restricted to their NH subsets or not, and the same was true when considering males alone. Studies in California, where over 40% of the Filipino COVID-19 deaths in 2020 occurred, have demonstrated that the population-level prevalence of chronic health conditions such as obesity, diabetes, and heart disease–risk factors for severe COVID-19 outcomes–are highest among Filipinos across all Asian subgroups [53–55]. While Filipino COVID-19 mortality rates were greater than that of Whites among males (whether comparisons were restricted to their NH subsets or not, and whether mortality rates were age-standardized or not), the same was not true for females. One potential contributing factor to this observed disjunction is that among Filipinos, male sex is associated with substantially greater prevalence of obesity [56], a risk factor for severe COVID-19 outcomes whose effect magnitude may be more pronounced among men [57, 58], but there is little to no sex difference in obesity prevalence among NH Whites [59]. Furthermore, greater occupational risks to SARS-CoV-2 exposure among Filipinos may also partially explain their greater relative COVID-19 mortality burden among Asians. For instance, Filipinos are vastly overrepresented in healthcare professions such as nursing [60], and approximately a quarter of Filipino adults are frontline healthcare workers [32]. As such, Filipinos were overrepresented among Asians who were providing direct medical care to COVID-19 patients during the early stages of the pandemic before vaccines were developed. Further compounding their increased risks of occupational exposure to SARS-CoV-2 in healthcare settings, Filipinos are more likely to live in multi-generational homes than Asians as a whole [61], making social distancing more difficult and increasing the likelihood of household SARS-CoV-2 transmission [62].

While COVID-19 was the third-leading cause of death in the U.S. for the year 2020 [63], deaths directly caused by COVID-19 are an incomplete measure of total pandemic-related mortality burden. Excess deaths–the difference between observed deaths and deaths expected in a counterfactual universe where the pandemic didn’t occur–is a more comprehensive metric to quantify the total impact of the COVID-19 pandemic on deaths, encompassing confirmed COVID-19 deaths, COVID-19 deaths among whom COVID-19 went undiagnosed, and deaths indirectly caused by COVID-19 (for example, deaths from pandemic-attributable treatment disruptions to chronic care management, or fatal drug overdoses resulting from the pandemic’s social and economic fallout). In fact, previous research has indicated that racial/ethnic disparities during the first year of the COVID-19 pandemic were greater for all-cause excess deaths than for COVID-19 deaths alone [64]. Future work characterizing the absolute and relative excess mortality burden for different Asian subgroups during the COVID-19 pandemic is warranted.

While there has been a dearth of data on population-level COVID-19 mortality outcomes for different Asian subgroups for much of the first two years of the pandemic, encouraging developments surrounding the CDC’s data environment have transpired. Beginning in December 2021, shortly before the final multiple cause of death data for 2020 were released, the CDC WONDER online data portal was enhanced to make provisional multiple cause of death data up to one month prior to the current month publicly available and downloadable, with the same degree of demographic and temporal granularity as the final multiple cause of death data that are released annually. This immensely positive development will more rapidly illuminate population-level disparities in mortality outcomes–especially those concerning Asian subgroups–whenever they may arise, elevating their attention and guiding data-driven public health efforts to direct resources to communities of greatest need.

Finally, a concurrent goal of this study was to bring greater awareness to the diverse Asian experiences during the COVID-19 pandemic in the U.S. through the rigorous quantitative analysis of data. During the COVID-19 pandemic and in the post-pandemic recovery, it is imperative not to overlook the needs of vulnerable Asian populations, such as Filipino men and Asian men outside of the six major Asian subgroups. Public health strategies to mitigate the effects of COVID-19 must avoid viewing Asians as a monolithic entity and must recognize the heterogeneous risk profiles within the U.S. Asian population across its diverse origin subgroups as well as by sex.

## Data Availability

This study used publicly available data that is referenced in the manuscript.

## Supplemental files

**File S1. Age-adjusted and sex-stratified COVID-19 mortality rates in 2020 for non-Hispanic Asians as a whole, non-Hispanic Asian subgroups, and non-Hispanic Whites for ages** < **45, ages 45–64, ages ≥ 65, ages ≥ 75, and all ages, and the corresponding mortality rate ratios and mortality rate differences relative to non-Hispanic Whites**.

**File S2. Age-adjusted and sex-stratified COVID-19 mortality rates in 2020 for Asians as a whole, Asian subgroups, and Whites for ages** < **45, ages 45–64, ages** ≥ **65, ages** ≥ **75, and all ages, and the corresponding mortality rate ratios and mortality rate differences relative to Whites**.

