## Supplementary material for "Variation in National COVID-19 Mortality Rates Across Asian Subgroups in the United States, 2020": File S1

**Table S1.1**: *Age-Adjusted and Sex-Stratified COVID-19 Mortality Rates in 2020 for Non-Hispanic Asians for Ages <45 Years, Ages 45-64 Years, Ages ≥75 Years, and All Ages and the Corresponding Mortality Rate Ratios and Mortality Rate Differences Relative to Non-Hispanic Whites*

| **Race/Ethnicity-Sex-Age Stratum** | **Deaths**[a] | **Mortality Rate**[a,b] (per 100,000) | **Mortality Rate Ratio**[a,b,c,d] | **Mortality Rate Difference**[a,b,c,d] (per 100,000) |
|---|---|---|---|---|
| **Non-Hispanic Asian** | | | | |
| **Male** | | | | |
| <45 years | 230 | 3.8 | 1.71 | 1.6 |
| 45-64 years | 1,628 | 67.0 | 1.60 | 25.1 |
| ≥65 years | 5,645 | 542.6 | 0.96 | -24.8 |
| ≥75 years | 3,637 | 806.6 | 0.85 | -147.8 |
| All ages | 7,503 | 87.1 | 1.04 | 3.5 |
| **Female** | | | | |
| <45 years | 114 | 1.7 | 1.19 | 0.3 |
| 45-64 years | 609 | 21.5 | 0.91 | -2.1 |
| ≥65 years | 4,467 | 362.2 | 0.85 | -62.1 |
| ≥75 years | 3,493 | 634.6 | 0.84 | -118.5 |
| All ages | 5,190 | 52.4 | 0.86 | -8.3 |

[a] 53 Asian male deaths and 29 Asian female deaths had missing Hispanic ethnicity and were excluded

[b] Age-standardized to the 2000 U.S. Standard Population

[c] 578 White male deaths and 289 White female deaths had missing Hispanic ethnicity and were excluded from the calculation

[d] 1 non-Hispanic White male death and 1 non-Hispanic White female death had missing ages and were excluded from the calculation

**Table S1.2**: *Age-Adjusted and Sex-Stratified COVID-19 Mortality Rates in 2020 for Non-Hispanic Asian Indians for Ages <45 Years, Ages 45-64 Years, Ages ≥75 Years, and All Ages and the Corresponding Mortality Rate Ratios and Mortality Rate Differences Relative to Non-Hispanic Whites*

| **Race/Ethnicity-Sex-Age Stratum** | **Deaths**[a] | **Mortality Rate**[a,b] (per 100,000) | **Mortality Rate Ratio**[a,b,c,d] | **Mortality Rate Difference**[a,b,c,d] (per 100,000) |
|---|---|---|---|---|
| **Non-Hispanic Asian Indian** | | | | |
| **Male** | | | | |
| <45 years | 34 | 1.5 | 0.69 | -0.7 |
| 45-64 years | 281 | 53.4 | 1.27 | 11.5 |
| ≥65 years | 904 | 573.1 | 1.01 | 5.7 |
| ≥75 years | 565 | 922.7 | 0.97 | -31.7 |
| All ages | 1,219 | 86.5 | 1.03 | 2.9 |
| **Female** | | | | |
| <45 years | 19 | 0.9 | 0.63 | -0.5 |
| 45-64 years | 68 | 14.5 | 0.62 | -9.1 |
| ≥65 years | 461 | 343.9 | 0.81 | -80.4 |
| ≥75 years | 317 | 568.0 | 0.75 | -185.1 |
| All ages | 548 | 47.9 | 0.79 | -12.7 |

[a] 8 Asian Indian male deaths and 7 Asian Indian female deaths had missing Hispanic ethnicity and were excluded

[b] Age-standardized to the 2000 U.S. Standard Population

[c] 578 White male deaths and 289 White female deaths had missing Hispanic ethnicity and were excluded from the calculation

[d] 1 non-Hispanic White male death and 1 non-Hispanic White female death had missing ages and were excluded from the calculation

**Table S1.3**: *Age-Adjusted and Sex-Stratified COVID-19 Mortality Rates in 2020 for Non-Hispanic Chinese for Ages <45 Years, Ages 45-64 Years, Ages ≥75 Years, and All Ages and the Corresponding Mortality Rate Ratios and Mortality Rate Differences Relative to Non-Hispanic Whites*

| Race/Ethnicity-Sex-Age Stratum | Deaths[a] | Mortality Rate[a,b] (per 100,000) | Mortality Rate Ratio[a,b,c,d] | Mortality Rate Difference[a,b,c,d] (per 100,000) |
|---|---|---|---|---|
| **Non-Hispanic Chinese** | | | | |
| **Male** | | | | |
| <45 years | 21 | 2.2 | 1.00 | 0.0 |
| 45-64 years | 198 | 37.3 | 0.89 | -4.6 |
| ≥65 years | 1,259 | 500.4 | 0.88 | -67.1 |
| ≥75 years | 932 | 807.3 | 0.85 | -147.1 |
| All ages | 1,478 | 74.0 | 0.88 | -9.6 |
| **Female** | | | | |
| <45 years | 9 | 0.7 | 0.50 | -0.7 |
| 45-64 years | 83 | 13.9 | 0.59 | -9.7 |
| ≥65 years | 1,009 | 424.0 | 1.00 | -0.2 |
| ≥75 years | 882 | 815.5 | 1.08 | 62.5 |
| All ages | 1,101 | 57.9 | 0.96 | -2.7 |

[a] 18 Chinese male deaths and 10 Chinese female deaths had missing Hispanic ethnicity and were excluded

[b] Age-standardized to the 2000 U.S. Standard Population

[c] 578 White male deaths and 289 White female deaths had missing Hispanic ethnicity and were excluded from the calculation

[d] 1 non-Hispanic White male death and 1 non-Hispanic White female death had missing ages and were excluded from the calculation

**Table S1.4**: *Age-Adjusted and Sex-Stratified COVID-19 Mortality Rates in 2020 for Non-Hispanic Filipinos for Ages <45 Years, Ages 45-64 Years, Ages ≥75 Years, and All Ages and the Corresponding Mortality Rate Ratios and Mortality Rate Differences Relative to Non-Hispanic Whites*

| Race/Ethnicity-Sex-Age Stratum | Deaths[a] | Mortality Rate[a,b] (per 100,000) | Mortality Rate Ratio[a,b,c,d] | Mortality Rate Difference[a,b,c,d] (per 100,000) |
|---|---|---|---|---|
| **Non-Hispanic Filipinos** | | | | |
| **Male** | | | | |
| <45 years | 51 | 9.2 | 4.09 | 6.9 |
| 45-64 years | 390 | 127.7 | 3.05 | 85.8 |
| ≥65 years | 1,221 | 712.0 | 1.25 | 144.6 |
| ≥75 years | 681 | 822.0 | 0.86 | -132.4 |
| All ages | 1,662 | 125.9 | 1.51 | 42.3 |
| **Female** | | | | |
| <45 years | 21 | 2.8 | 1.98 | 1.4 |
| 45-64 years | 216 | 41.9 | 1.77 | 18.3 |
| ≥65 years | 997 | 384.0 | 0.91 | -40.3 |
| ≥75 years | 695 | 626.9 | 0.83 | -126.2 |
| All ages | 1,234 | 60.5 | 1.00 | -0.1 |

[a] 12 Filipino male deaths and 4 Filipino female deaths had missing Hispanic ethnicity and were excluded

[b] Age-standardized to the 2000 U.S. Standard Population

[c] 578 White male deaths and 289 White female deaths had missing Hispanic ethnicity and were excluded from the calculation

[d] 1 non-Hispanic White male death and 1 non-Hispanic White female death had missing ages and were excluded from the calculation

**Table S1.5**: *Age-Adjusted and Sex-Stratified COVID-19 Mortality Rates in 2020 for Non-Hispanic Japanese for Ages <45 Years, Ages 45-64 Years, Ages ≥75 Years, and All Ages and the Corresponding Mortality Rate Ratios and Mortality Rate Differences Relative to Non-Hispanic Whites*

| Race/Ethnicity-Sex-Age Stratum | Deaths[a] | Mortality Rate[a,b] (per 100,000) | Mortality Rate Ratio[a,b,c,d] | Mortality Rate Difference[a,b,c,d] (per 100,000) |
|---|---|---|---|---|
| **Non-Hispanic Japanese** | | | | |
| **Male** | | | | |
| <45 years | 1 | 0.6 | 0.25 | -1.7 |
| 45-64 years | 31 | 28.9 | 0.69 | -12.9 |
| ≥65 years | 202 | 236.7 | 0.42 | -330.7 |
| ≥75 years | 163 | 413.4 | 0.43 | -541.0 |
| All ages | 234 | 37.2 | 0.45 | -46.4 |
| **Female** | | | | |
| <45 years | 1 | 0.7 | 0.46 | -0.8 |
| 45-64 years | 3 | 1.6 | 0.07 | -22.0 |
| ≥65 years | 369 | 203.1 | 0.48 | -221.1 |
| ≥75 years | 344 | 385.2 | 0.51 | -367.9 |
| All ages | 373 | 26.8 | 0.44 | -33.8 |

[a] 2 Japanese male deaths had missing Hispanic ethnicity and were excluded

[b] Age-standardized to the 2000 U.S. Standard Population

[c] 578 White male deaths and 289 White female deaths had missing Hispanic ethnicity and were excluded from the calculation

[d] 1 non-Hispanic White male death and 1 non-Hispanic White female death had missing ages and were excluded from the calculation

**Table S1.6**: *Age-Adjusted and Sex-Stratified COVID-19 Mortality Rates in 2020 for Non-Hispanic Koreans for Ages <45 Years, Ages 45-64 Years, Ages ≥75 Years, and All Ages and the Corresponding Mortality Rate Ratios and Mortality Rate Differences Relative to Non-Hispanic Whites*

| Race/Ethnicity-Sex-Age Stratum | Deaths[a] | Mortality Rate[a,b] (per 100,000) | Mortality Rate Ratio[a,b,c,d] | Mortality Rate Difference[a,b,c,d] (per 100,000) |
|---|---|---|---|---|
| **Non-Hispanic Korean** | | | | |
| **Male** | | | | |
| <45 years | 7 | 1.8 | 0.82 | -0.4 |
| 45-64 years | 54 | 25.7 | 0.61 | -16.2 |
| ≥65 years | 492 | 471.0 | 0.83 | -96.4 |
| ≥75 years | 390 | 777.5 | 0.81 | -176.9 |
| All ages | 553 | 67.3 | 0.81 | -16.3 |
| **Female** | | | | |
| <45 years | 4 | 0.7 | 0.46 | -0.8 |
| 45-64 years | 28 | 9.7 | 0.41 | -13.9 |
| ≥65 years | 572 | 501.8 | 1.18 | 77.5 |
| ≥75 years | 521 | 983.9 | 1.31 | 230.8 |
| All ages | 604 | 66.9 | 1.10 | 6.3 |

[a] 1 Korean male death and 1 Korean female death had missing Hispanic ethnicity and were excluded

[b] Age-standardized to the 2000 U.S. Standard Population

[c] 578 White male deaths and 289 White female deaths had missing Hispanic ethnicity and were excluded from the calculation

[d] 1 non-Hispanic White male death and 1 non-Hispanic White female death had missing ages and were excluded from the calculation

**Table S1.7**: *Age-Adjusted and Sex-Stratified COVID-19 Mortality Rates in 2020 for Non-Hispanic Vietnamese for Ages <45 Years, Ages 45-64 Years, Ages ≥75 Years, and All Ages and the Corresponding Mortality Rate Ratios and Mortality Rate Differences Relative to Non-Hispanic Whites*

| **Race/Ethnicity-Sex-Age Stratum** | **Deaths**[a] | **Mortality Rate**[a,b] (per 100,000) | **Mortality Rate Ratio**[a,b,c,d] | **Mortality Rate Difference**[a,b,c,d] (per 100,000) |
|---|---|---|---|---|
| **Non-Hispanic Vietnamese** | | | | |
| **Male** | | | | |
| <45 years | 20 | 6.0 | 2.69 | 3.8 |
| 45-64 years | 169 | 78.4 | 1.87 | 36.5 |
| ≥65 years | 641 | 452.0 | 0.80 | -115.5 |
| ≥75 years | 431 | 734.1 | 0.77 | -220.3 |
| All ages | 830 | 79.5 | 0.95 | -4.1 |
| **Female** | | | | |
| <45 years | 9 | 1.8 | 1.23 | 0.3 |
| 45-64 years | 43 | 15.0 | 0.64 | -8.6 |
| ≥65 years | 445 | 333.7 | 0.79 | -90.5 |
| ≥75 years | 347 | 607.0 | 0.81 | -146.1 |
| All ages | 497 | 47.3 | 0.78 | -13.3 |

[a] 3 Vietnamese male deaths had missing Hispanic ethnicity and were excluded

[b] Age-standardized to the 2000 U.S. Standard Population

[c] 578 White male deaths and 289 White female deaths had missing Hispanic ethnicity and were excluded from the calculation

[d] 1 non-Hispanic White male death and 1 non-Hispanic White female death had missing ages and were excluded from the calculation

**Table S1.8**: *Age-Adjusted and Sex-Stratified COVID-19 Mortality Rates in 2020 for Non-Hispanic Other Asians for Ages <45 Years, Ages 45-64 Years, Ages ≥75 Years, and All Ages and the Corresponding Mortality Rate Ratios and Mortality Rate Differences Relative to Non-Hispanic Whites*

| **Race/Ethnicity-Sex-Age Stratum** | **Deaths**[a] | **Mortality Rate**[a,b] (per 100,000) | **Mortality Rate Ratio**[a,b,c,d] | **Mortality Rate Difference**[a,b,c,d] (per 100,000) |
|---|---|---|---|---|
| **Non-Hispanic Other Asians** | | | | |
| **Male** | | | | |
| <45 years | 96 | 7.3 | 3.28 | 5.1 |
| 45-64 years | 505 | 111.0 | 2.65 | 69.1 |
| ≥65 years | 926 | 838.7 | 1.48 | 271.3 |
| ≥75 years | 475 | 1,073.5 | 1.12 | 119.1 |
| All ages | 1,527 | 137.2 | 1.64 | 53.6 |
| **Female** | | | | |
| <45 years | 51 | 4.2 | 2.90 | 2.7 |
| 45-64 years | 168 | 38.9 | 1.65 | 15.3 |
| ≥65 years | 614 | 414.9 | 0.98 | -9.3 |
| ≥75 years | 387 | 595.8 | 0.79 | -157.2 |
| All ages | 833 | 64.6 | 1.07 | 4.0 |

[a] 9 Other Asian male deaths and 7 Other Asian female deaths had missing Hispanic ethnicity and were excluded

[b] Age-standardized to the 2000 U.S. Standard Population

[c] 578 White male deaths and 289 White female deaths had missing Hispanic ethnicity and were excluded from the calculation

[d] 1 non-Hispanic White male death and 1 non-Hispanic White female death had missing ages and were excluded from the calculation

**Table S1.9**: *Age-Adjusted and Sex-Stratified COVID-19 Mortality Rates in 2020 for Non-Hispanic Whites for Ages <45 Years, Ages 45-64 Years, Ages ≥75 Years, and All Ages*

| Race/Ethnicity-Sex-Age Stratum | Deaths[a,b] | Mortality Rate[a,b,c] (per 100,000) | Mortality Rate Ratio | Mortality Rate Difference (per 100,000) |
|---|---|---|---|---|
| **Non-Hispanic Whites** | | | | |
| **Male** | | | | |
| <45 years | 1,092 | 2.2 | Reference | Reference |
| 45-64 years | 13,173 | 41.9 | Reference | Reference |
| ≥65 years | 95,792 | 567.4 | Reference | Reference |
| ≥75 years | 71,316 | 954.4 | Reference | Reference |
| All ages | 110,058 | 83.6 | Reference | Reference |
| **Female** | | | | |
| <45 years | 690 | 1.4 | Reference | Reference |
| 45-64 years | 7,770 | 23.6 | Reference | Reference |
| ≥65 years | 90,619 | 424.2 | Reference | Reference |
| ≥75 years | 75,451 | 753.1 | Reference | Reference |
| All ages | 99,080 | 60.6 | Reference | Reference |

[a] 578 White male deaths and 289 White female deaths had missing Hispanic ethnicity and were excluded

[b] 1 non-Hispanic White male death and 1 non-Hispanic White female death had missing ages and were excluded

[c] Age-standardized to the 2000 U.S. Standard Population
