## Supplementary material for "Variation in National COVID-19 Mortality Rates Across Asian Subgroups in the United States, 2020": File S2

**Table S2.1**: *Age-Adjusted and Sex-Stratified COVID-19 Mortality Rates in 2020 for Asians for Ages <45 Years, Ages 45-64 Years, Ages ≥75 Years, and All Ages and the Corresponding Mortality Rate Ratios and Mortality Rate Differences Relative to Whites*

| Race-Sex-Age Stratum | Deaths | Mortality Rate[a] (per 100,000) | Mortality Rate Ratio[a,b] | Mortality Rate Difference[a,b] (per 100,000) |
|---|---|---|---|---|
| **Asian** | | | | |
| **Male** | | | | |
| <45 years | 236 | 3.8 | 0.62 | -2.4 |
| 45-64 years | 1,666 | 66.9 | 0.88 | -9.4 |
| ≥65 years | 5,721 | 540.3 | 0.85 | -98.5 |
| ≥75 years | 3,688 | 807.1 | 0.78 | -228.9 |
| All ages | 7,623 | 86.8 | 0.84 | -16.3 |
| **Female** | | | | |
| <45 years | 115 | 1.7 | 0.61 | -1.1 |
| 45-64 years | 622 | 21.6 | 0.61 | -13.8 |
| ≥65 years | 4,524 | 360.2 | 0.79 | -98.6 |
| ≥75 years | 3,531 | 630.2 | 0.79 | -165.2 |
| All ages | 5,261 | 52.1 | 0.76 | -16.4 |

[a] Age-standardized to the 2000 U.S. Standard Population

[b] 1 White male death and 1 White female death had missing ages and were excluded

**Table S2.2**: *Age-Adjusted and Sex-Stratified COVID-19 Mortality Rates in 2020 for Asian Indians for Ages <45 Years, Ages 45-64 Years, Ages ≥75 Years, and All Ages and the Corresponding Mortality Rate Ratios and Mortality Rate Differences Relative to Whites*

| Race-Sex-Age Stratum | Deaths | Mortality Rate[a] (per 100,000) | Mortality Rate Ratio[a,b] | Mortality Rate Difference[a,b] (per 100,000) |
|---|---|---|---|---|
| **Asian Indian** | | | | |
| **Male** | | | | |
| <45 years | 34 | 1.5 | 0.25 | -4.6 |
| 45-64 years | 284 | 53.7 | 0.70 | -22.6 |
| ≥65 years | 911 | 570.3 | 0.89 | -68.6 |
| ≥75 years | 569 | 929.4 | 0.90 | -106.5 |
| All ages | 1,229 | 86.2 | 0.84 | -16.9 |
| **Female** | | | | |
| <45 years | 19 | 0.9 | 0.33 | -1.8 |
| 45-64 years | 71 | 15.1 | 0.43 | -20.3 |
| ≥65 years | 469 | 349.7 | 0.76 | -109.2 |
| ≥75 years | 324 | 580.5 | 0.73 | -214.9 |
| All ages | 559 | 48.8 | 0.71 | -19.8 |

[a] Age-standardized to the 2000 U.S. Standard Population

[b] 1 White male death and 1 White female death had missing ages and were excluded

**Table S2.3**: *Age-Adjusted and Sex-Stratified COVID-19 Mortality Rates in 2020 for Chinese for Ages <45 Years, Ages 45-64 Years, Ages ≥75 Years, and All Ages and the Corresponding Mortality Rate Ratios and Mortality Rate Differences Relative to Whites*

| Race-Sex-Age Stratum | Deaths | Mortality Rate[a] (per 100,000) | Mortality Rate Ratio[a,b] | Mortality Rate Difference[a,b] (per 100,000) |
|---|---|---|---|---|
| **Chinese[e]** | | | | |
| **Male** | | | | |
| <45 years | 21 | 2.2 | 0.36 | -4.0 |
| 45-64 years | 204 | 37.0 | 0.49 | -39.3 |
| ≥65 years | 1,274 | 497.6 | 0.78 | -141.2 |
| ≥75 years | 944 | 804.7 | 0.78 | -231.2 |
| All ages | 1,499 | 73.5 | 0.71 | -29.5 |
| **Female** | | | | |
| <45 years | 9 | 0.7 | 0.26 | -2.0 |
| 45-64 years | 84 | 13.8 | 0.39 | -21.6 |
| ≥65 years | 1,021 | 426.7 | 0.93 | -32.1 |
| ≥75 years | 892 | 821.8 | 1.03 | 26.4 |
| All ages | 1,114 | 58.3 | 0.85 | -10.3 |

[a] Age-standardized to the 2000 U.S. Standard Population

[b] 1 White male death and 1 White female death had missing ages and were excluded

**Table S2.4**: *Age-Adjusted and Sex-Stratified COVID-19 Mortality Rates in 2020 for Filipinos for Ages <45 Years, Ages 45-64 Years, Ages ≥75 Years, and All Ages and the Corresponding Mortality Rate Ratios and Mortality Rate Differences Relative to Whites*

| Race-Sex-Age Stratum | Deaths | Mortality Rate[a] (per 100,000) | Mortality Rate Ratio[a,b] | Mortality Rate Difference[a,b] (per 100,000) |
|---|---|---|---|---|
| **Filipinos** | | | | |
| **Male** | | | | |
| <45 years | 52 | 8.4 | 1.36 | 2.2 |
| 45-64 years | 404 | 128.4 | 1.68 | 52.1 |
| ≥65 years | 1,262 | 731.6 | 1.15 | 92.7 |
| ≥75 years | 706 | 850.9 | 0.82 | -185.0 |
| All ages | 1,718 | 128.1 | 1.24 | 25.0 |
| **Female** | | | | |
| <45 years | 21 | 2.7 | 0.97 | -0.1 |
| 45-64 years | 221 | 41.8 | 1.18 | 6.3 |
| ≥65 years | 1,022 | 379.2 | 0.83 | -79.7 |
| ≥75 years | 709 | 615.4 | 0.77 | -180.0 |
| All ages | 1,264 | 59.7 | 0.87 | -8.8 |

[a] Age-standardized to the 2000 U.S. Standard Population

[b] 1 White male death and 1 White female death had missing ages and were excluded

**Table S2.5**: *Age-Adjusted and Sex-Stratified COVID-19 Mortality Rates in 2020 for Japanese for Ages <45 Years, Ages 45-64 Years, Ages ≥75 Years, and All Ages and the Corresponding Mortality Rate Ratios and Mortality Rate Differences Relative to Whites*

| Race-Sex-Age Stratum | Deaths | Mortality Rate[a] (per 100,000) | Mortality Rate Ratio[a,b] | Mortality Rate Difference[a,b] (per 100,000) |
|---|---|---|---|---|
| **Japanese** | | | | |
| **Male** | | | | |
| <45 years | 3 | 1.3 | 0.21 | -4.9 |
| 45-64 years | 32 | 28.4 | 0.37 | -47.9 |
| ≥65 years | 202 | 230.9 | 0.36 | -407.9 |
| ≥75 years | 163 | 413.4 | 0.40 | -622.6 |
| All ages | 237 | 36.8 | 0.36 | -66.2 |
| **Female** | | | | |
| <45 years | 1 | 0.6 | 0.21 | -2.2 |
| 45-64 years | 3 | 1.5 | 0.04 | -33.9 |
| ≥65 years | 370 | 199.9 | 0.44 | -259.0 |
| ≥75 years | 345 | 379.4 | 0.48 | -415.9 |
| All ages | 374 | 26.3 | 0.38 | -42.2 |

[a] Age-standardized to the 2000 U.S. Standard Population

[b] 1 White male death and 1 White female death had missing ages and were excluded

**Table S2.6**: *Age-Adjusted and Sex-Stratified COVID-19 Mortality Rates in 2020 for Koreans for Ages <45 Years, Ages 45-64 Years, Ages ≥75 Years, and All Ages and the Corresponding Mortality Rate Ratios and Mortality Rate Differences Relative to Whites*

| Race/Ethnicity-Sex-Age Stratum | Deaths | Mortality Rate[a] (per 100,000) | Mortality Rate Ratio[a,b] | Mortality Rate Difference[a,b] (per 100,000) |
|---|---|---|---|---|
| **Korean** | | | | |
| **Male** | | | | |
| <45 years | 7 | 1.8 | 0.30 | -4.3 |
| 45-64 years | 55 | 26.2 | 0.34 | -50.1 |
| ≥65 years | 492 | 471.0 | 0.74 | -167.8 |
| ≥75 years | 390 | 777.5 | 0.75 | -258.5 |
| All ages | 554 | 67.5 | 0.65 | -35.6 |
| **Female** | | | | |
| <45 years | 4 | 0.6 | 0.23 | -2.1 |
| 45-64 years | 28 | 9.7 | 0.27 | -25.8 |
| ≥65 years | 573 | 502.7 | 1.10 | 43.8 |
| ≥75 years | 522 | 985.9 | 1.24 | 190.5 |
| All ages | 605 | 67.0 | 0.98 | -1.6 |

[a] Age-standardized to the 2000 U.S. Standard Population

[b] 1 White male death and 1 White female death had missing ages and were excluded

**Table S2.7**: *Age-Adjusted and Sex-Stratified COVID-19 Mortality Rates in 2020 for Vietnamese for Ages <45 Years, Ages 45-64 Years, Ages ≥75 Years, and All Ages and the Corresponding Mortality Rate Ratios and Mortality Rate Differences Relative to Whites*

| **Race/Ethnicity-Sex-Age Stratum** | **Deaths** | **Mortality Rate**[a] (per 100,000) | **Mortality Rate Ratio**[a,b] | **Mortality Rate Difference**[a,b] (per 100,000) |
|---|---|---|---|---|
| **Vietnamese** | | | | |
| **Male** | | | | |
| <45 years | 20 | 6.0 | 0.97 | -0.2 |
| 45-64 years | 170 | 78.9 | 1.03 | 2.6 |
| ≥65 years | 643 | 448.4 | 0.70 | -190.4 |
| ≥75 years | 433 | 726.6 | 0.70 | -309.3 |
| All ages | 833 | 79.1 | 0.77 | -24.0 |
| **Female** | | | | |
| <45 years | 9 | 1.7 | 0.63 | -1.0 |
| 45-64 years | 43 | 15.0 | 0.42 | -20.4 |
| ≥65 years | 445 | 333.7 | 0.73 | -125.2 |
| ≥75 years | 347 | 607.0 | 0.76 | -188.3 |
| All ages | 497 | 47.3 | 0.69 | -21.3 |

[a] Age-standardized to the 2000 U.S. Standard Population

[b] 1 White male death and 1 White female death had missing ages and were excluded

**Table S2.8**: *Age-Adjusted and Sex-Stratified COVID-19 Mortality Rates in 2020 for Other Asians for Ages <45 Years, Ages 45-64 Years, Ages ≥75 Years, and All Ages and the Corresponding Mortality Rate Ratios and Mortality Rate Differences Relative to Whites*

| Race-Sex-Age Stratum | Deaths | Mortality Rate[a] (per 100,000) | Mortality Rate Ratio[a,b] | Mortality Rate Difference[a,b] (per 100,000) |
|---|---|---|---|---|
| **Other Asians** | | | | |
| **Male** | | | | |
| <45 years | 99 | 7.4 | 1.21 | 1.3 |
| 45-64 years | 517 | 112.1 | 1.47 | 35.7 |
| ≥65 years | 937 | 809.1 | 1.27 | 170.3 |
| ≥75 years | 483 | 1,007.0 | 0.97 | -29.0 |
| All ages | 1,553 | 133.7 | 1.30 | 30.7 |
| **Female** | | | | |
| <45 years | 52 | 4.2 | 1.53 | 1.5 |
| 45-64 years | 172 | 39.1 | 1.10 | 3.7 |
| ≥65 years | 624 | 395.3 | 0.86 | -63.6 |
| ≥75 years | 392 | 548.7 | 0.69 | -246.7 |
| All ages | 848 | 62.2 | 0.91 | -6.4 |

[a] Age-standardized to the 2000 U.S. Standard Population

[b] 1 White male death and 1 White female death had missing ages and were excluded

**Table S2.9**: *Age-Adjusted and Sex-Stratified COVID-19 Mortality Rates in 2020 for Whites for Ages <45 Years, Ages 45-64 Years, Ages ≥75 Years, and All Ages*

| **Race-Sex-Age Stratum** | **Deaths** | **Mortality Rate**[a,b] (per 100,000) | **Mortality Rate Ratio** | **Mortality Rate Difference** (per 100,000) |
|---|---|---|---|---|
| **Whites** | | | | |
| **Male** | | | | |
| <45 years | 3,989 | 6.2 | Reference | Reference |
| 45-64 years | 27,073 | 76.3 | Reference | Reference |
| ≥65 years | 119,442 | 638.8 | Reference | Reference |
| ≥75 years | 84,540 | 1,036.0 | Reference | Reference |
| All ages | 150,505 | 103.0 | Reference | Reference |
| **Female** | | | | |
| <45 years | 1,755 | 2.7 | Reference | Reference |
| 45-64 years | 13,430 | 35.4 | Reference | Reference |
| ≥65 years | 107,967 | 458.9 | Reference | Reference |
| ≥75 years | 87,060 | 795.3 | Reference | Reference |
| All ages | 123,153 | 68.6 | Reference | Reference |

[a] Age-standardized to the 2000 U.S. Standard Population

[b] 1 White male death and 1 White female death had missing ages and were excluded
